# Higher polygenic risk for melanoma is associated with improved survival

**DOI:** 10.1101/2022.03.07.22272003

**Authors:** Mathias Seviiri, Richard A. Scolyer, D. Timothy Bishop, Mark M. Iles, Serigne N. Lo, Johnathan R. Stretch, Robyn P.M. Saw, Omgo E. Nieweg, Kerwin F. Shannon, Andrew J. Spillane, Scott D. Gordon, Catherine M. Olsen, David C. Whiteman, Maria T. Landi, John F. Thompson, Georgina V. Long, Stuart MacGregor, Matthew H. Law

## Abstract

**Background:** Although there are well-known prognostic factors for survival from cutaneous melanoma (CM) such as primary tumour thickness and stage of the tumour at diagnosis, the role of germline genetic factors in determining survival is not well understood.

**Objective:** To perform a genome-wide association study (GWAS) meta-analysis of melanoma-specific survival (MSS), and test whether a CM-susceptibility polygenic risk score (PRS) is associated with MSS.

**Methods:** We conducted two Cox proportional-hazard GWAS of MSS using data from the Melanoma Institute Australia (MIA; 5,762 patients with melanoma; 800 deaths from melanoma) and UK Biobank (UKB: 5,220 patients with melanoma; 241 deaths from melanoma). The GWAS were adjusted for age, sex and the first ten genetic principal components, and combined in a fixed-effects inverse-variance-weighted meta-analysis. Significant (P<5×10^−8^) results were investigated in the Leeds Melanoma Cohort (LMC; 1,947 patients with melanoma; 370 melanoma deaths). We also developed a CM-susceptibility PRS using a large independent GWAS meta-analysis (23,913 cases, 342,870 controls). The PRS was tested for an association with MSS in the MIA and UKB cohorts, with replication in the LMC.

**Results:** Two loci were significantly associated with MSS in the meta-analysis of MIA and UKB with lead SNPs rs41309643 (G allele frequency 1.6%, hazard ratio [HR] 2.09, 95% confidence interval [CI] 1.61-2.71, P=2.08×10^−8^) on chromosome 1, and rs75682113 (C allele frequency 1.8%, HR=2.38, 95% CI=1.77—3.21, P=1.07×10^−8^) on chromosome 7. While neither SNP replicated (P>0.05) in the LMC, rs75682113 was significantly associated in the combined discovery and replication sets and requires confirmation in additional cohorts.

After adjusting for age at diagnosis, sex and the first ten principal components, a one standard deviation increase in the CM-susceptibility PRS was associated with improved MSS in the discovery meta-analysis (HR=0.88, 95% CI=0.83—0.94, P=6.93×10^−5^; I^2^=88%). The association with the PRS was not replicated (P > 0.05) in LMC, but remained significantly associated with MSS in the meta-analysis of the discovery and replication results.

**Conclusion:** We found two loci potentially associated with MSS, and evidence that increased germline genetic susceptibility to develop CM may be associated with improved MSS.

## INTRODUCTION

Cutaneous melanoma (CM) is the third most common skin cancer and is responsible for over 1,300 deaths in Australia annually (Cancer Australia 2019) and more than 7,000 deaths in the United States of America (USA) (NCI 2021). While survival rates have been improving since 2013, likely due to advances in immunotherapies and BRAF-targeted therapies, management of CM remains a major public health burden, with an annual cost of over AUD 200 million in Australia and USD 24 billion in the US (Elliott et al. 2017; Zaorsky et al. 2021).

CM-susceptibility is driven by host factors including skin pigmentation and number of naevi, as well as environmental factors, most importantly exposure to ultraviolet radiation (Gandini, Sera, Cattaruzza, Pasquini, Abeni, et al. 2005; Gandini, Sera, Cattaruzza, Pasquini, Zanetti, et al. 2005; Mitra et al. 2012; Y.-M. Chang et al. 2009; Veierød et al. 2010). Germline genetic factors can influence the risk of developing CM through modification of these host risk factors, and other biological pathways; genome-wide association studies (GWAS) have identified over 50 CM-susceptibility loci (Landi et al. 2020).

Although there are well known prognostic factors for melanoma-specific survival (MSS) including primary tumour thickness, ulceration, mitotic rate, melanoma type, anatomical site and the stage of the tumour at diagnosis (Cherobin et al. 2018; Kibrité et al. 2016), the role of host genetic factors in MSS is not well understood. Death of a relative from CM is associated with poorer MSS, raising the possibility that germline genetic factors influence survival (Brandt, Sundquist, and Hemminki 2011). Higher naevus count has been associated with improved survival (Ribero et al. 2015). Naevus count is strongly influenced by germline genetics (Zhu et al. 1999; Wachsmuth et al. 2001), and is the strongest risk factor for the development of melanoma (Olsen, Carroll, and Whiteman 2010), suggesting germline genetic risk for CM may also impact survival. Telomere length is another biological pathway to high genetic CM-susceptibility (Burke et al. 2013) and may also influence MSS (Rachakonda et al. 2018).

A powerful approach to test whether germline genetic risk for a given disease or trait (e.g. risk for CM) influences another trait (e.g. MSS) is to combine individual genetic effects in a polygenic risk score (PRS). Death from all causes has been associated with the joint effect of PRSs associated with risk of a range of diseases (e.g. coronary artery disease, pancreatic cancer, and lung cancer) or associated with mortality risk factors (e.g. cholesterol, sleep duration) (Meisner et al. 2020), suggesting that germline risk for development of a disease can help predict outcomes. However, it is not known whether a genetic predisposition to CM influences melanoma outcomes.

To explore these two questions, we first aimed to identify germline genetic factors that influence MSS by performing a large-scale GWAS of MSS. Following this we assessed whether a PRS for CM-susceptibility (referred to as PRS_susceptibility) was associated with MSS.

## METHODS AND MATERIALS

### Genome-wide association studies of melanoma-specific survival

#### Discovery cohort 1: Melanoma Institute Australia

Samples for this cohort were derived from the Melanoma Institute Australia (MIA) Biospecimen Bank (protocol HREC/10/RPAH/530) and patient information from the MIA Research Database (protocol HREC/11/RPAH/444). With written, informed consent, patients with histo-pathological confirmed CM cases managed at MIA, Sydney, Australia were identified from this Biospecimen Bank and Database. Participants’ clinical and biospecimen data were captured and prospectively collected follow-up for outcomes including death due to melanoma. MIA study protocols were approved by the Sydney Local Health District Ethics Review Committee, Royal Prince Alfred Hospital, Camperdown, Australia. Participants were genotyped in phases using the Oncoarray in 2014 and 2016, and the Global Screening Array in 2018 (Illumina, San Diego).

Full details of the GWAS data cleaning quality control for both MIA datasets have been previously reported (Landi et al. 2020; Liyanage et al. 2021). Briefly, for Oncoarray genotyped samples, individuals were removed based on high genotype missingness (> 3%), extreme heterozygosity (± 0.05 from the mean), being related to other samples (identified by descent pihat > 0.15), or were more than 6 standard deviations (SDs) from the means of principal components (PCs) 1 and 2 of a European reference population (Landi et al. 2020). In addition, single nucleotide polymorphisms (SNPs) were removed if they were missing more than 3% of their calls, had a minor allele frequency (MAF) < 0.01, or their Hardy–Weinberg equilibrium (HWE) P-value was less than 5 × 10^−10^ for patients with melanoma or less than 5 × 10^−4^ in CM-free individuals in Landi et al., (Landi et al. 2020). Individuals genotyped on the Global Screening Array were removed due to high genotype missingness (> 5%), non-European ancestry or relatedness (as above), and SNPs were excluded due to a low MAF (< 0.01), high missingness (> 5%), HWE P < 1 × 10^−6^, or a low GenTrain score (< 0.6) (Liyanage et al. 2021). The cleaned genotyped data were batched by their genotyping array (Oncoarray and Global Screening Array) and imputed to the Haplotype Reference Consortium (v1) panel using the University of Michigan imputation server (Loh et al. 2016).

For this study, the primary endpoint was MSS which was ascertained through MIA clinical records and linkage to Australian Cancer Registries (including the New South Wales Cancer Registry), electoral rolls, and the Birth and Death Register. This analysis was restricted to 5,672 participants of European ancestry diagnosed with CM. For participants with multiple CM, the first primary CM was used to define the start point. MSS survival time (in years) was defined as the duration between the date of diagnosis of the (first) primary CM, and the date of death due to melanoma. Patients were censored on the last day of follow-up or when they died of non-melanoma causes.

#### Discovery cohort 2: UK Biobank

UK Biobank (UKB) is a large population-based cohort of approximately 500,000 adult participants (40-70 years at recruitment) recruited with informed consent from the United Kingdom between 2006 and 2010. Participants were followed up for disease outcomes including death from melanoma. Details on participant recruitment, phenotype measurement and genotyping have been published elsewhere (Sudlow et al. 2015; Bycroft et al. 2018). In brief, participants were genotyped using the UK Biobank Axiom Array and the UK BiLEVE Axiom Array (Affymetrix Inc, California, USA) and imputed using the Haplotype Reference Consortium and UK10K reference panels. The study was approved by the United Kingdom’s National North West Multi-Centre Research Ethics Committee. For this present study, we included 5,220 participants of European ancestry with histo-pathologically confirmed invasive CM based on the International Classification of Diseases (ICD) 10 (UKB data field 40006) and 9 (data field 40013) and ICD for Oncology, 3rd edition codes (data field 40011) for melanoma. Participants were then filtered for missingness (<3%), relatedness (identity by descent pihat < 0.2), and population ancestry outliers (from the European reference). The primary endpoint was MSS which was ascertained through linkage of the participant records with Cancer Registries, electoral rolls, and the Birth and Death Register in the UK.

#### Replication cohort: Leeds Melanoma Cohort

The Leeds Melanoma Cohort (LMC) is a population-based cohort of 2,184 participants diagnosed with incident melanoma between September 2000 and December 2012 and residing in Yorkshire and the North of England (Newton-Bishop et al. 2015). Details on the recruitment, follow-up and phenotype/genotype data processing have been published previously (Newton-Bishop et al. 2015, 2010; Bishop et al. 2009). In brief, for two periods (September 2000 - December 2001, and July 2003 to December 2005) recruitment was restricted to patients with a primary tumour thickness of > 0.75 mm, while all patients with invasive melanoma were invited to participate between January 2002 and June 2003, and between January 2006 - 2012. Melanoma survival information was collected by direct communication with patients and their families, clinical records and from national registers.

Melanoma diagnoses were clinico-histopathologically confirmed through data linkage with the Northern and Yorkshire Cancer Registry and Information Service. Samples were genotyped using the Infinium HumanOmniExpressExome array (Illumina San Diego, CA, USA). After genetic quality control procedures (filtering for missingness, relatedness, and population outliers), this present study was restricted to 1,947 participants with genetic and phenotype data, and consent. Ethical approval for research involving the LMC was obtained from the Northern and Yorkshire Research Ethics Committee, and all participants provided written informed consent.

SNPs with MAF < 0.03, control Hardy-Weinberg equilibrium (HWE) P < 10^−4^ or missingness > 0.03 were excluded, as were any individuals with call rates <0.97, identified as first degree relatives and/or European outliers by principal components analysis. Samples were imputed using the Haplotype Reference Consortium panel at the University of Michigan imputation server (Loh et al. 2016) and variants with an imputation quality score <0.5 or MAF<0.0001 were discarded.

#### Statistical analysis: Genome-wide association study of melanoma-specific survival

First, we conducted two GWAS of MSS in the MIA cohort (5,762 patients with melanoma and 800 melanoma-specific deaths) and in UKB cohort (5,220 melanoma patients and 241 melanoma-specific deaths). Using Cox proportional-hazard modelling, hazard ratios (HRs) were computed using PLINK 1.9 (C. C. Chang et al. 2015) and the R *survival* package (Therneau 2020). In both the MIA and UKB analyses, we adjusted for age, sex and the first ten PCs; in the MIA cohort we also adjusted for genotyping batch. Analysis was restricted to participants of European ancestry and SNPs with MAF > 0.5%, and an imputation quality score > 0.5.

Next, we conducted a meta-analysis for both GWAS (N=10,982 and 1,041 melanoma deaths) using a fixed-effects inverse-variance weighted model in METAL (Willer, Li, and Abecasis 2010). In addition, measures of heterogeneity (such as I^2^) were computed. Lead genome-wide significant (P < 5 × 10^−8^) SNPs independent at linkage disequilibrium (LD) r^2^ < 0.1 were identified using FUMA v1.3.6a (https://fuma.ctglab.nl/) (Watanabe et al. 2017).

Lead SNPs were tested for replication in the LMC (N=1,947 patients with melanoma and 370 melanomas-specific deaths). The replication p-value threshold was set to 0.05. Next, we conducted a fixed- and random-effects inverse-variance meta-analysis of the lead SNPs from all three sets (MIA, UKB and LMC) using METAL (Willer, Li, and Abecasis 2010). For the two lead SNPs the nearest gene, and any significant expression quantitative trait loci (eQTLs) were identified using FUMA v1.3.6a (Watanabe et al. 2017).

### Cutaneous melanoma polygenic risk score

#### Cutaneous melanoma risk discovery cohorts and GWAS meta-analysis

As the three MSS GWAS cohorts contributed to the discovery CM-susceptibility GWAS meta-analysis (Landi et al. 2020), and overlap between datasets used to generate, optimise or test PRS can lead to overfitting and other biases (Lambert, Abraham, and Inouye 2019), we re-analysed the CM-susceptibility GWAS meta-analysis excluding the three MSS GWAS datasets. We further excluded the QSkin Sun and Health Study cohort to use as an independent data set to validate the generated PRSs. Details on recruitment, case definitions, genotyping, quality control, imputation approaches and ethical approvals for each cohort have been extensively described before (Landi et al. 2020). The updated meta-analysis consisted of 23,913 cases, and 342,870 controls of European ancestry from Europe, Australia and the United States of America (USA) (**Supplementary Table 1**).

With the exception of the self-reported 23andMe, Inc. dataset, all CM cases were histopathologically confirmed; previous work has shown that 23andMe cases are very similar to the confirmed cases: the susceptibility loci show very similar effects in the self-reported and confirmed CM cases (Landi et al. 2020). Each study was approved by the human research ethics committee at their respective institution, and all participants provided written informed consent. Specifically, for 23andMe, participants provided written informed consent and participated in the research online, under a protocol approved by the external AAHRPP-accredited IRB, Ethical & Independent Review Services.

Only SNPs with an imputation quality score > 0.5 were included, and a fixed-effects inverse variance weighted meta-analysis of log odds ratios (ORs) was performed using PLINK 1.9 (C. C. Chang et al. 2015). Next, we selected 6,342,711 non-ambiguous, autosomal, bi-allelic GWAS meta-analysis SNPs with a MAF > 1% that were present in the validation (QSkin) and target (MIA and UKB) cohorts, and in the LD reference panel.

#### CM PRS validation cohort: The QSkin Sun and Health Study cohort

The QSkin Sun and Health Study (QSkin) cohort is a population-based cohort comprising over 43,000 adult participants recruited from Queensland, Australia. Detailed information on participant recruitment, phenotype measurement, genotyping and quality control measures have been published elsewhere (Olsen et al. 2012; Landi et al. 2020). In summary, 18,087 participants were genotyped using the Global Screening Array [Illumina, San Diego, USA], and individuals were removed if they had non-European ancestry (6 s.d from the mean of PC1 and PC2 of 1000 Genomes European samples), were related to another participant (one from each pair removed if identity by descent pihat value > 0.1875), or had high genotype missingness (> 3%). SNPs were also removed due to HWE violations (P < 1 × 10^−6^), a low GenTrain score (< 0.6), or a low MAF (< 0.01). Cleaned genotype data were imputed to the haplotype reference consortium (v1) panel using the University of Michigan imputation server (Loh et al. 2016).

The Human Research Ethics Committee of QIMR Berghofer Medical Research Institute, Brisbane, Australia approved the study protocol and all participants provided written informed consent. We selected 16,708 participants (1,285 histopathologically confirmed CM cases and 15,423 controls) of European ancestry. CM cases were ascertained through data linkage with the Queensland Cancer Registry as well as assessing histopathology reports from pathology laboratories in Queensland.

#### Generation of the cutaneous melanoma polygenic risk score models

We used the CM-susceptibility GWAS data (generated above) and an LD reference panel of 2,000 unrelated individuals of European ancestry from UKB, to generate 30 PRS_susceptibility models at 1 megabase (Mb), 2 Mb, 3 Mb, 4 Mb and 5 Mb of LD radii each with varying fractions of causal SNPs i.e. 1 (F0), 0.1 (F1), 0.01 (F2), 0.001 (F3), 0.0001 (F4), and 0.00002 (F5). For this analysis we used LDpred, a Bayesian method that utilises all SNPs in the discovery GWAS (here CM-susceptibility GWAS), and their LD information, to derive LD-adjusted effect estimates (log ORs) for the trait (here CM-susceptibility) (Vilhjálmsson et al. 2015).

#### Validation of the cutaneous melanoma polygenic risk score in QSkin cohort

Next, we used the QSkin validation cohort to select the optimally performing PRS. Next, for each model we computed scores for 16,708 individuals (1,285 melanoma cases and 15,423 controls) in the QSkin Cohort using the LDpred-adjusted effect sizes (log ORs) and the imputed allelic dosages using PLINK 1.9 (C. C. Chang et al. 2015). Then we computed and used Nagelkerke’s R^2^ (Nagelkerke 1991) to select the optimally performing PRS_susceptibility model by comparing the model fit for CM risk ∼ PRS_susceptibility +age + sex +10 PCs, and a null model (CM risk ∼ age + sex +10 PCs) using the PredictABEL R package (Kundu et al. 2011). Model performances are presented in **Figure 1**, and the best performing PRS model was used in the subsequent analyses.

**Figure 1:**
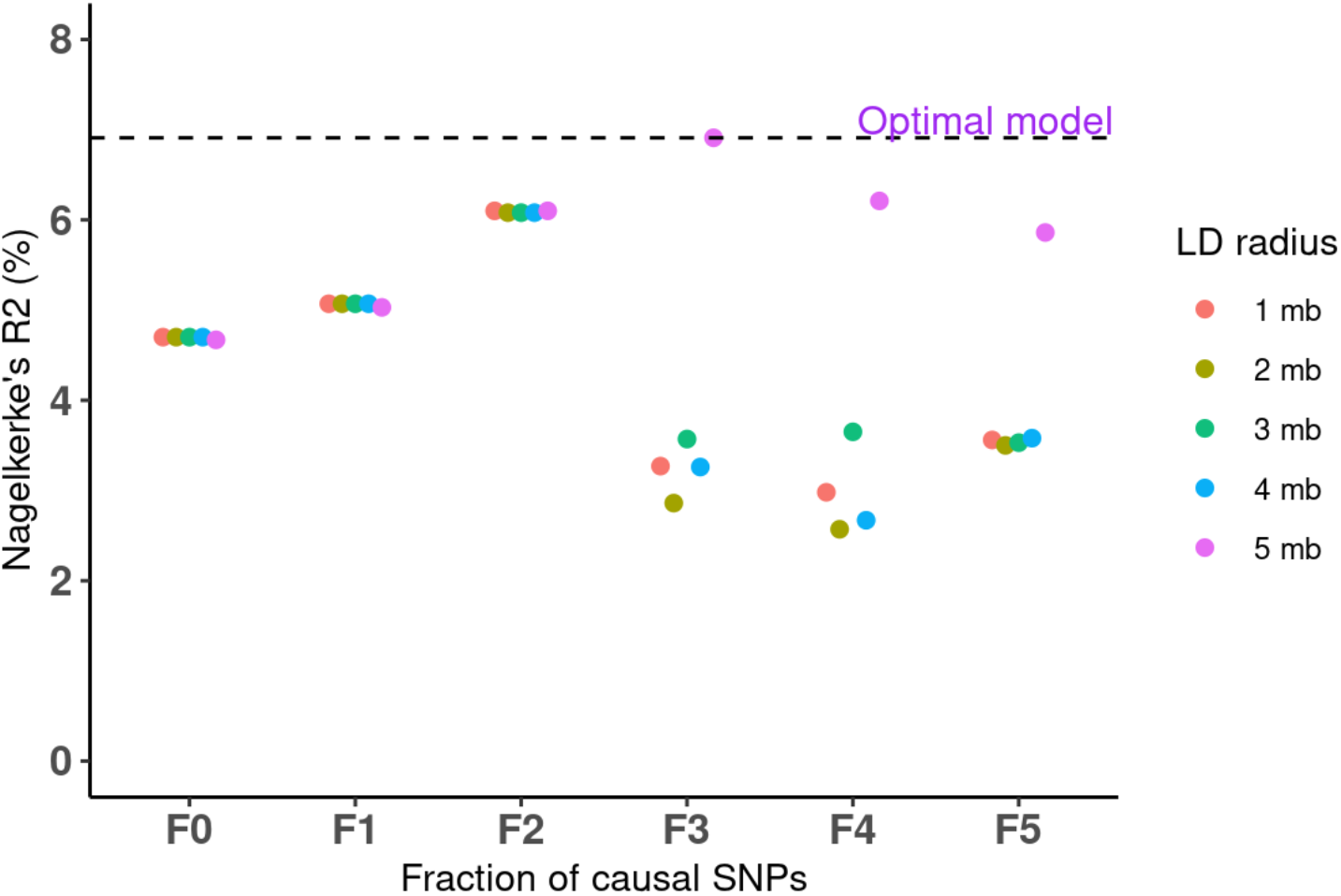
Cutaneous melanoma polygenic risk score model performance in the validation cohort (QSkin). *The x-axis represents the different melanoma* polygenic risk score (PRS) *modelling varying fractions of causal SNPs, 1 (F0), 0.1 (F1), 0.01 (F2), 0.001 (F3), 0.0001 (F4) and 0.00002 (F5), and differing linkage disequilibrium (LD) radii, 1 megabase (Mb), 2 Mb, 3 Mb, 4 Mb and 5 Mb. The y-axis represents Nagelkerke’s R*^*2*^ *(%) for each of the 30 PRS models. The horizontal dashed black line highlights the optimal model (F3 and 5Mb) with the highest Nagelkerke’s R*^*2*^ *of 7.03%*.

#### Testing for association between cutaneous melanoma polygenic risk scores and melanoma-specific survival

The best performing PRS_susceptibility model was applied to the MIA and UKB cohorts using imputed allelic dosages and PLINK 1.9. The PRS was normalised to have a mean of 0 and an SD of 1 and tested for association with MSS in a Cox proportional hazard model adjusted for age at diagnosis, sex and the first ten PCs using the *survival* package in R (Therneau 2020). We further calculated the MSS HR and 95% confidence interval (CI) per SD increase in the PRS_susceptibility. Next, we conducted a fixed- and random-effects inverse-variance meta-analysis to compute the pooled HR and 95% CI using the meta R package (Balduzzi, Rücker, and Schwarzer 2019). We then we tested for association between MSS and the same PRS_susceptibility in the LMC, adjusting for the same covariates. Finally, we meta-analysed the MIA, UKB and LMC results.

#### Sensitivity analyses for polygenic susceptibility to melanoma and melanoma-specific survival

Pigmentation and naevus count loci are major biological pathways for CM-susceptibility (Duffy et al. 2018; Landi et al. 2020). We further explored whether any association between the PRS_susceptibility and MSS was driven by SNPs associated with pigmentation and/or naevi pathways (**Supplementary Information**). In addition, we generated PRSs for pigmentation (PRS_P_), naevus count (PRS_N_) and telomere length (PRS_TL_) and tested whether they were associated with MSS (**Supplementary Information**). To rule out the possibility of thin or slow-growing melanomas influencing the PRS-survival association, we explored the potential influence of tumour stage, thickness and lead-time bias on any associations (**Supplementary Information**).

## RESULTS

### Baseline characteristics of the melanoma survival cohorts

This analysis was restricted to 5,762 melanoma patients in the MIA cohort, 5,220 in the UKB cohort, and 1,947 in the LMC. Summary data on mean age at diagnosis, sex, duration of follow up and the number of melanoma-specific deaths are presented in **Table 1**.

**Table 1:** Characteristics of Melanoma Institute Australia, UK Biobank and Leeds Melanoma Cohorts.

| Characteristic | MIA | UKB | LMC |
| --- | --- | --- | --- |
| Number | 5762 | 5220 | 1947 |
| Mean age in years (SD) | 60.1 (15.4) | 56.78 (11.2) | 55.05 (13.4) |
| Number of males (%) | 3,478 (60.4) | 2,231 (42.7) | 839 (43.1) |
| Mean duration of follow up in years (SD) | 5.82 (6.4) | 13.69 (8.7) | 7.29 (3.7) |
| Number of melanoma-specific deaths (%) | 800 (13.9) | 241 (4.6) | 370 (19.0) |

*MIA = Melanoma Institute Australia cohort, UKB = UK Biobank, LMC = Leeds Melanoma Cohort*
*SD - Standard deviation, N - Number, % - percent*

### Genome-wide significant genetic variants for melanoma-specific survival

A MSS GWAS meta-analysis of the MIA and UKB cohorts identified two independent genome-wide significant (P < 5 × 10^−8^) loci **(Table 2**, and **Supplementary Figure 1);** rs41309643 (P = 2.08 × 10^−8^) on chromosome 1 (1q42.13) and rs75682113 (P = 1.07 × 10^−8^) on chromosome 7 (7p14.1) (**Table 2**). However, neither SNP was replicated in the LMC (rs41309643 P = 0.679 and rs75682113 P = 0.411 (**Table 2**, and **Supplementary Table 2)**. Following the meta-analysis of all three cohorts, rs41309643 was no longer formally significant at P < 5 × 10^−8^ (HR = 1.83, 95% CI = 1.45—2.30, P = 3.21 × 10^−7^) with high heterogeneity metrics (**Table 2**). rs75682113 remained genome-wide significant with no significant evidence of heterogeneity (C-allele HR = 2.23, 95% CI = 1.68—2.95, P = 2.13 × 10^−8^**; Table 2**).

**Table 2:**
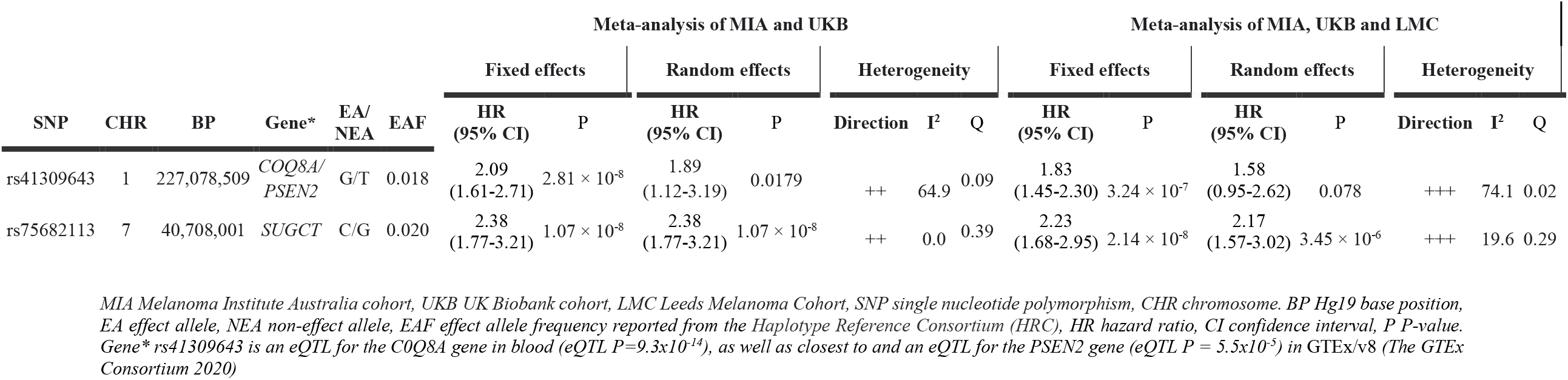
Genetic variants for melanoma-specific survival in the discovery cohorts (MIA+UKB) and replication cohort (LMC).

Rs41309643 on chromosome 1 is an intron of the *PSEN2* gene and is associated with the expression of the *Coenzyme Q8A* (*COQ8A*) (formerly *ADCK3*) gene in blood. *COQ8A* is induced by *p53* in response to DNA damage and inhibition of *COQ8A* counteracts p53-induced apoptosis (Iiizumi et al. 2002). rs75682113 on chromosome 7 is in an intron of the *Succinyl-Coa:Glutarate-Coa Transferase* (*SUGCT*) gene. This SNP has not been reported as an *eQTL* for any genes. Independent variants in the *SUGCT* gene have been associated with glutaric aciduria type 3 disease susceptibility (Sherman et al. 2008).

### The optimal cutaneous melanoma susceptibility polygenic risk score model

Of the thirty PRS tested, the model with the F3 causal fraction (0.001) and a 5 Mb LD radius performed best, with a Nagelkerke’s R^2^ of 7.02% (**Figure 1**), and was used in all subsequent analyses.

### Association of polygenic susceptibility to melanoma and melanoma-specific survival

After adjusting for age at diagnosis, sex and the first ten PCs, a one SD increase in the PRS_susceptibility was associated with improved MSS in a fixed-effects meta-analysis of MIA and UKB cohorts (HR = 0.88, 95% CI = 0.83—0.94, P = 6.93 × 10^−5^). However, the association between the PRS_susceptibility and MSS was highly heterogeneous across the two studies (I^2^ = 87.7%, 95% CI = 52.4—96.8%). Although not statistically significant, the magnitude and direction for the random effects model was also consistent with the fixed-effects results (fixed effects model HR = 0.92, 95% CI = 0.75—1.13, P = 0.43). The inverse association between polygenic susceptibility to melanoma and MSS persisted after excluding genomic regions associated with naevus count (fixed-effects HR = 0.91, 95% CI = 0.86—0.97, P =0.0038; random-effects HR = 0.93, 95% CI = 0.83—1.03, P = 0.16) and pigmentation (fixed-effects HR = 0.91, 95% CI = 0.850—.97, P =0.0023; random-effects HR = 0.93 95% CI = 0.82—1.06, P = 0.26). The association between polygenic risk for melanoma and MSS was not replicated (P > 0.05) in the LMC; however, the directions of the effect estimates were consistent (**Figure 2**).

**Figure 2:**
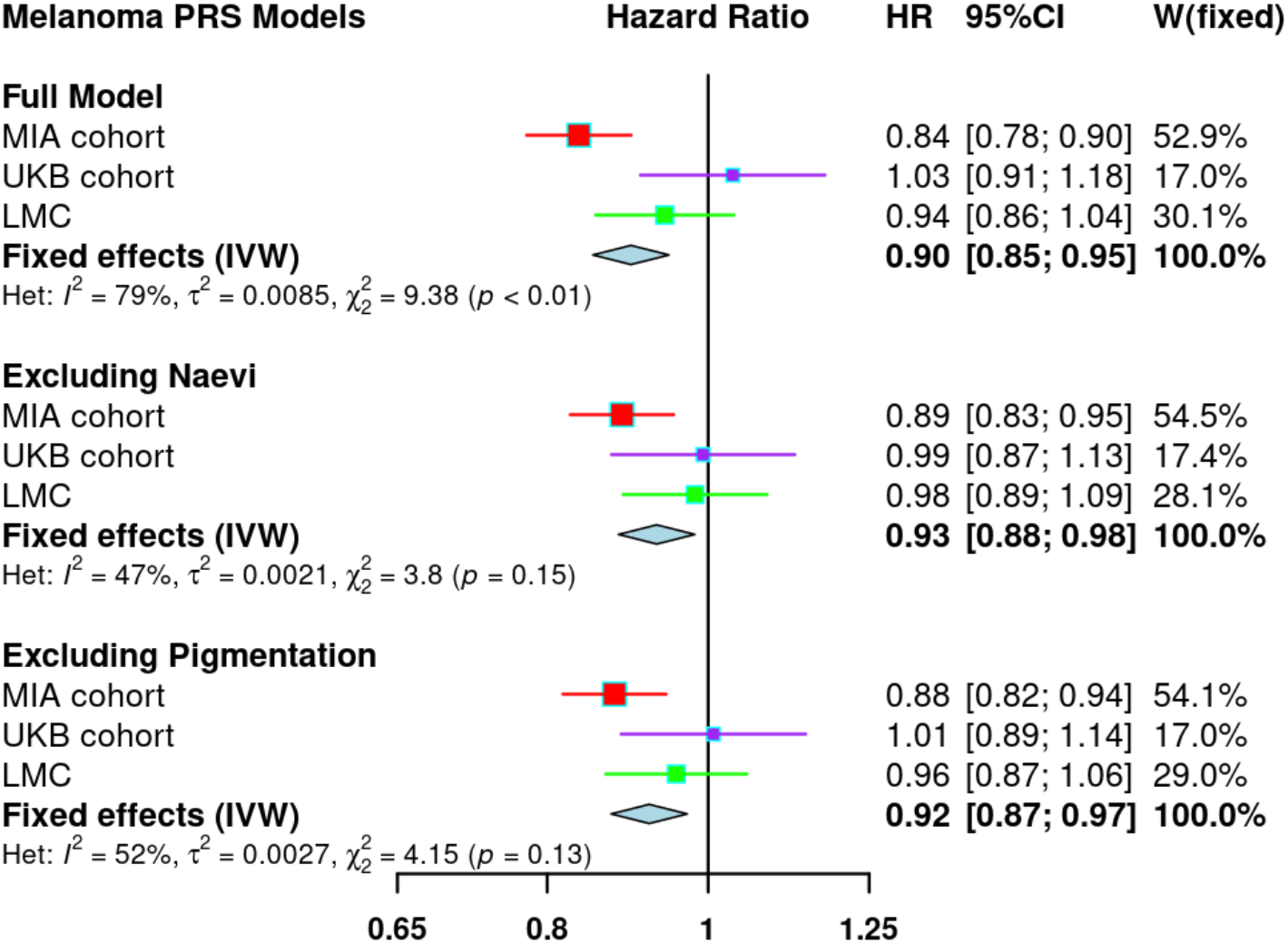
Association of polygenic risk for cutaneous melanoma and melanoma-specific survival. All models were adjusted for age, sex and the first 10 PCs and additionally genotype batch effects in the MIA analysis. HRs were estimated using Cox proportional-hazards models. The full model refers to the PRS_susceptibility (CM PRS), while for the remaining two models the PRS_susceptibility respectively excluded SNPs in the naevus count and pigmentation pathways. MIA-Melanoma Institute Australia, UKB - United Kingdom Biobank, LMC - Leeds Melanoma Cohort, IVW-Inverse variance weighted methods, Het-heterogeneity, HR-hazard ratio. CI-confidence interval.

In a meta-analysis of the three cohorts a one SD increase in the PRS_susceptibility was still associated with improved MSS (fixed-effects HR = 0.90, 95% CI = 0.85—0.95, P = 6.35 × 10^−5^ ; random-effects HR = 0.93, 95% CI = 0.83—1.04, P = 0.20), even after excluding naevus and pigmentation loci (**Figure 2**). There was substantial heterogeneity across the three studies (I^2^ = 78.7%, 95% CI = 31.6— 93.4%). Sensitivity analyses showed that the skin colour PRS was also associated with improved MSS (PRS_**P**_; HR = 0.90, 95% CI = 0.85—0.96, P = 1.1 × 10^−3^), while the naevus count PRS also provided suggestive evidence (PRS_**N**_; HR = 0.95, 95%CI = 0.89—1.02, P = 0.179) (**Supplementary Figure 5**).

### Influence from melanoma prognostic factors and lead-time bias in the MIA Cohort

In the MIA cohort the PRS_susceptibility remained associated with improved survival after excluding participants with melanoma *in-situ*, and those with an unknown stage (HR = 0.84, 95% CI = 0.78— 0.90, P = 2.15 × 10^−6^). In addition, the association was consistent even after adjusting for age, sex, 10 PCs, AJCC 2010 Stage, and primary tumour thickness (HR = 0.84, 95% CI = 0.78—0.91, P = 1.90 × 10^−5^) (**Table 3**). There was also no evidence for interaction by the tumour stage or tumour thickness **(Table 3**). In a stratified analysis, there was no evidence that the association between the PRS and MSS differed by tumour stage (**Figure 3a**) and primary tumour thickness at diagnosis (**Figure 3b**). The PRS_TL_ was suggestive but not significantly associated with MSS in the MIA cohort (PRS_TL_; HR = 0.90, 95% CI = 0.64 — 1.27, P = 0.5504). After excluding the first two years of follow-up (following diagnosis), there was no evidence of lead-time bias (survival bias) (HR = 0.84, 95% CI = 0.77—0.91, P = 4.03 × 10^−5^).

**Table 3:**
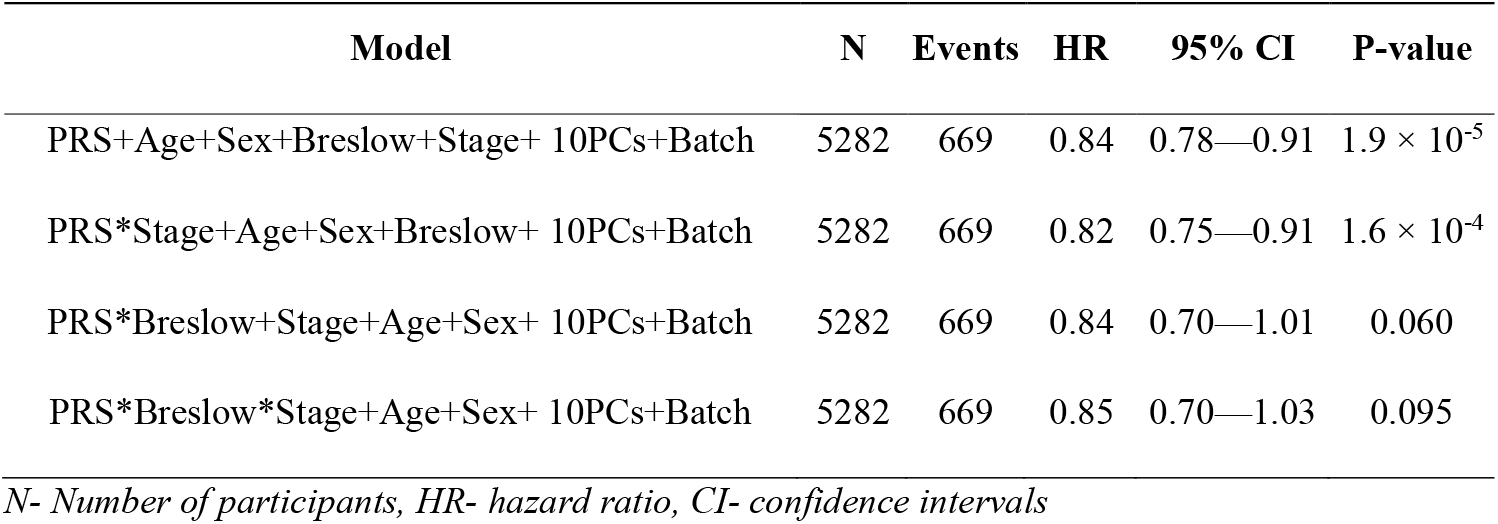
**Testing for an interaction between the polygenic susceptibility to melanoma and survival prognostic factors in the MIA Cohort**

| Model | N | Events | HR | 95% CI | P-value |
| --- | --- | --- | --- | --- | --- |
| PRS+Age+Sex+Breslow+Stage+ 10PCs+Batch | 5282 | 669 | 0.84 | 0.78—0.91 | $1.9 \times 10^{-5}$ |
| PRS*Stage+Age+Sex+Breslow+ 10PCs+Batch | 5282 | 669 | 0.82 | 0.75—0.91 | $1.6 \times 10^{-4}$ |
| PRS*Breslow+Stage+Age+Sex+ 10PCs+Batch | 5282 | 669 | 0.84 | 0.70—1.01 | 0.060 |
| PRS*Breslow*Stage+Age+Sex+ 10PCs+Batch | 5282 | 669 | 0.85 | 0.70—1.03 | 0.095 |

*N- Number of participants, HR- hazard ratio, CI- confidence intervals*

**Figure 3:**
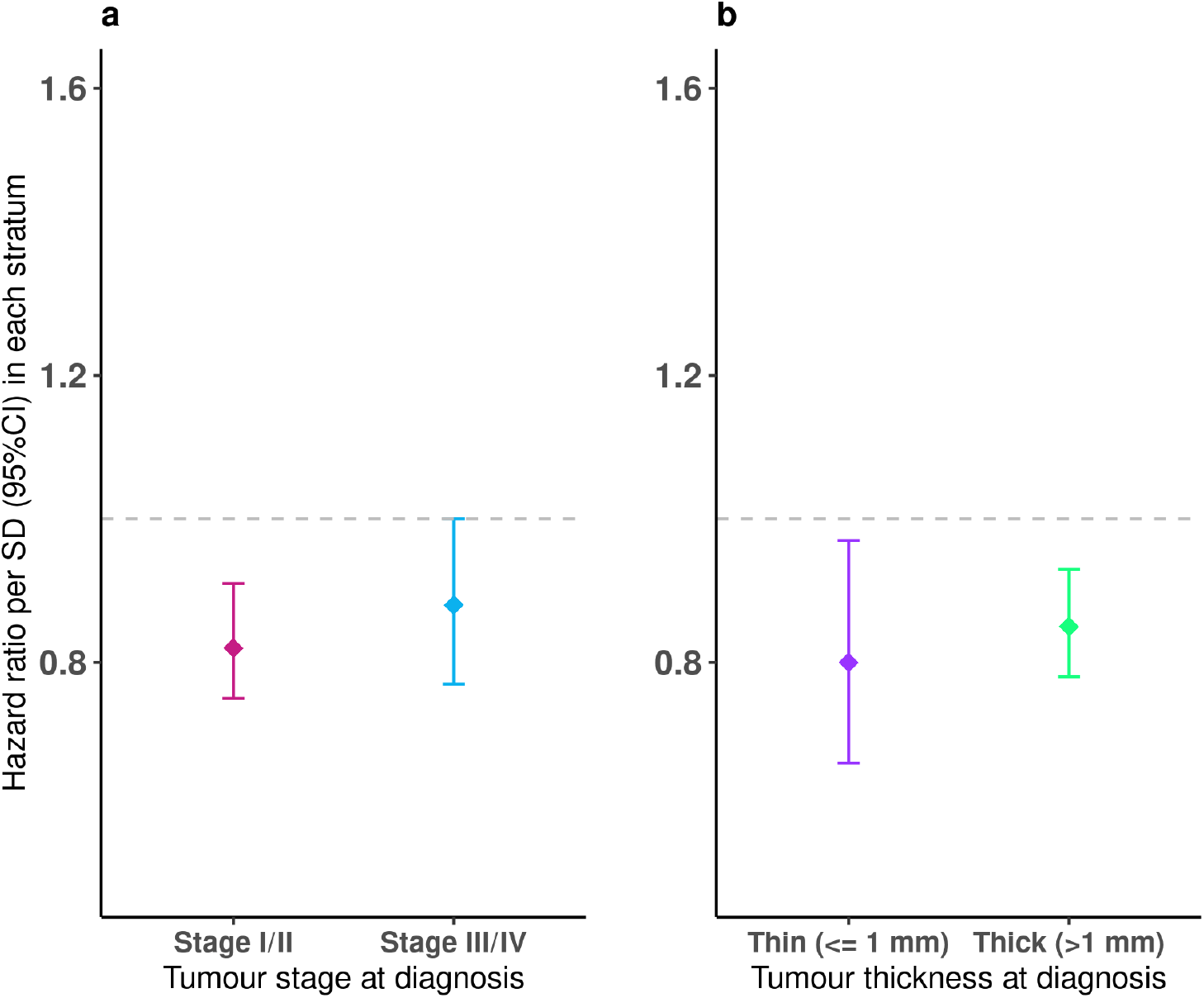
Stratified analysis of the PRS and MSS association by the AAJC Stage and primary tumour thickness in the MIA Cohort. The y-axis represents the hazard ratio for MSS per standard deviation (SD) increase in the PRS. Error bars show the 95% confidence interval of the HR. The x-axis shows the strata for tumour stage and thickness at diagnosis of melanoma. The dashed grey line represents a null effect at a hazard ratio of 1. **Panel 3a** shows the association between the CM PRS and MSS stratified by the AJCC 2010 tumour stage, after controlling for age at diagnosis, sex, the first 10 ancestral components and genotype batch effects. Stage I/II included 4493 participants and 427 melanoma deaths, while stage III/IV included 789 participants and 242 melanoma deaths. **Panel 3b** reports the association of the CM PRS and MSS stratified by the primary tumour thickness, after controlling for age at diagnosis, sex, the first 10 ancestral components and genotype batch effects. The thin (<= 1 mm) stratum included 1,898 participants and 122 melanoma deaths, while the thick (> 1 mm) stratum included 3,384 participants and 547 melanoma deaths.

## DISCUSSION

In this study, we performed the largest GWAS for MSS to date using data from Australia and the United Kingdom and potentially have identified two independent, novel, genome-wide significant (P < 5×10^−8^) loci for MSS at 1q42.13 and 7p14.1. While the two loci did not formally replicate in an independent cohort, the confidence intervals (particularly for rs75682113) in the replication set overlap the estimate from the discovery cohorts. Confirmation of these two loci will require replication in larger cohorts. rs75682113 is particularly promising as it was genome-wide significant (P < 5× 10^−8^) in our meta-analysis of the discovery and replication samples.

In addition, we report evidence that increased genetic susceptibility for CM, as measured by a one SD increase in a PRS_susceptibility, was significantly associated with improved MSS. However, caution is required as the result was primarily driven by a strong association in the MIA cohort. Genetic susceptibility to CM is primarily driven by loci in the pigmentation and naevus count pathways (Cust et al. 2018). HRs for PRS_susceptibility and MSS were slightly attenuated (but still with a significant association) when we removed SNPs in either pathway. In turn PRS designed specifically for these traits were also associated (though not significantly for naevus count) with MSS. In addition, the PRS for telomere length (another pathway to both CM susceptibility and survival) was not significantly associated with MSS in our sensitivity analysis. These pathway-analysis results suggest that if genetic propensity to CM is associated with improved survival it is not simply due to pigmentation, nevus count or telomere length.

However, this study suggests that if there is a true association, its magnitude may differ across populations, presumably due to environmental and other effects. Firstly, the MIA and UKB meta-analysis results did not replicate in the LMC. Secondly, the high heterogeneity metrics (e.g. I^2^) indicates that the effect sizes may not be consistent across the three studies, with a very strong result in the MIA cohort and weaker associations in the UK samples (**Table 1 and Supplementary Table 2**). Although the fixed-effects model shows a strong statistically significant association, the results are not significant for the random-effects model even when they are of a similar magnitude. The observed heterogeneity may be due to differences in recruitment, where the MIA cohort recruitment was from clinics as opposed to the population-based UKB and LMC. It is also possible that the strong inverse result in Australia is influenced by overdiagnosis for melanoma (Welch, Mazer, and Adamson 2021). It is estimated that 54% of all melanomas and 15% invasive melanomas in Australia are over-diagnosed (Glasziou, Bell, and Barratt 2020). Thus, patients may be diagnosed with non-lethal melanoma and subsequently exhibit improved survival. However, recent evidence suggests that regular skin checks (which may lead to overdiagnosis for melanoma) are not associated with MSS (Watts et al. 2021). Since sun exposure is associated with improved MSS (Berwick et al. 2005; Rosso et al. 2008), it is also possible that differences in high or long-term sun and ultraviolet-radiation exposure in Australia are in part responsible for the heterogeneity.

In a more detailed analysis in the MIA cohort, our study suggests that this inverse association is consistent even after further adjusting for (and testing for interaction with) strong predictors of MSS like tumour stage and primary tumour thickness at diagnosis. The stratified analysis shows that the association is not modified by primary tumour thickness or stage. Thus, if replicated in additional cohorts, a CM-susceptibility PRS is potentially an independent prognostic factor for MSS.

To our knowledge, while no prior study has examined the association of a CM susceptibility PRS and survival outcome, similar inverse relationships have been found in other cancers e.g. higher breast cancer PRSs and better breast cancer prognosis/characteristics (Holm et al. 2016; Li et al. 2018). Also, a follicular lymphoma PRS was associated with improved overall survival among women in a population in the USA (Zhong et al. 2020). BRCA1/2 mutations which increase breast cancer risk were associated with better overall survival among triple-negative breast cancer women (Baretta et al. 2016). A CAD PRS was inversely associated with all-cause mortality (OR=0.91; 95% CI=0.85-0.98), and ischaemic stroke (OR = 0.78; 95% CI=0.67-0.90) in CAD patients (Howe et al. 2020).

The mechanisms underlying this inverse association are unclear. Particularly for MSS, it could be that a higher genetic risk for CM leads to thin melanomas or slow-growing melanomas that are less lethal (Adami et al. 2017; Halpern and Marghoob 2004; Maurichi et al. 2014), and respond better to treatment. However, detailed analysis in the MIA cohort showed no difference in survival for both thin and thick tumour categories. In addition, after excluding the initial two years of follow up, the results were consistent, suggesting there is no survival/ lead-time bias.

As noted in our study above, higher nevus counts may be associated with a lower chance of dying from melanoma (Ribero et al. 2015). It is possible however that those with large numbers of naevi are subjected to increased screening, which may lead to overdiagnosis and greater survival relative to those with fewer moles (Autier et al. 2015). However, as already indicated, increased screening is not associated with MSS (Watts et al. 2021).

Another possible mechanism could be via gene-environment interaction, where those at highest genetic risk of CM benefit more from treatment (e.g immunotherapy), as it is the case for those at high genetic risk for coronary artery disease (CAD) and treatment benefits from PCSK9 inhibitors in the FOURIER and ODYSSEY OUTCOMES trials (Marston et al. 2020; Damask et al. 2020).

### Potential clinical utility

This study presents new insights that highlight the potential clinical utility of *PRS_susceptibility* for profiling and monitoring patients for melanoma outcomes following diagnosis during the “melanoma follow-up care program” (Trotter et al. 2013; Farma and Abdulla 2015). In combination with other prognostic factors, it could be used to guide patient care e.g. counselling on modification of mortality-related non-genetic behaviours and lifestyle factors, or guide the direction of patient-specific treatment to help improve survival after diagnosis. It may also be useful for the stratification of patients while recruiting into clinical trials evaluating melanoma treatment and outcomes.

## Conclusions

In a GWAS meta-analysis of MSS, we identified two novel loci potentially associated with survival from cutaneous melanoma, both of which contain candidate genes linked to tumour progression; however, replication in large independent cohorts is required. In line with observations in other cancers and complex diseases, increased germline genetic susceptibility for CM was strongly but heterogeneously associated with improved MSS. If validated, a PRS_susceptibility could be used to predict melanoma outcomes after diagnosis and profile patients for personalised care.

## Data Availability Statement

The pruning and thresholding (P+T) versions of polygenic risk scores CM can be accessed at the polygenic risk score catalogue (https://www.pgscatalog.org/) upon publication. CM GWAS summary statistics used to generate the LDPred PRSs can be accessed as indicated by Landi et al 2020. Underlying data for the cohorts used in the paper are available through application to the respective cohorts; UKB (http://www.ukbiobank.ac.uk/wp-content/uploads/2012/09/Access-Procedures-2011-1.pdf); MIA (https://www.melanoma.org.au/research/collaborate-on-research-with-mia/); Q-Skin (By application to Q-Skin Principal Investigator David Whiteman).

## Author Contributions

Conceptualization: MS, GVL, RAS, MHL, SM; Data collection, generation, and curation: MS, MHL, SDG, MTL, DCW, CMO, SM, SNL, JRS, RPMS, OEN, KFS, AJS, JFT; Genotyping and imputation: MTL, SM, SDG, MMI, DTB, MHL. Formal analysis: MS, MHL, DTB; Funding acquisition: SM, MHL, DCW, GVL, RAS, DTB, MTL; Investigation: MS, MHL, GVL, RAS, DCW, CMO, SM; Methodology: MS, RAS, GVL, MHL, SM; Project administration: MS, MHL, SM; Resources: MHL, GVL, RAS, MTL, DCW, SM; Software: MS; Supervision: SM, MHL; Visualisation: MS; Writing - original draft preparation: MS. Writing - subsequent drafts preparation: MS, SM, MHL, VJ, HB, DCW, CMO, SDG, GVL, RAS, MTL, DCW, MMI, DTB, SNL, JRS, RPMS, OEN, KFS, AJS, JFT.

All authors contributed to the final version of the manuscript.

## ABBREVIATIONS

(AJCC): American Joint Committee on Cancer
(CM): cutaneous melanoma
(GWAS): genome-wide association studies
(HR): hazard ratio
(ICD): International Classification of Diseases
(Mb): megabase
(LD): linkage disequilibrium
(MIA): Melanoma Institute Australia
(MSS): melanoma-specific survival
(MAF): minor allele frequency
(PRS): polygenic risk score
(PCs): principal components
(QSkin): QSkin Sun and Health Study
(SNP): single nucleotide polymorphism
(SD): standard deviation
(UKB): UK Biobank
(R^2^): variance
(LMC): Leeds Melanoma Cohort
(CI): and Confidence interval

## QIMR BERGHOFER MEDICAL RESEARCH INSTITUTE

The study was supported by a program grant (APP1073898) and a project grant (APP1063061) from the Australian National Health and Medical Research Council (NHMRC). SM and DCW are supported by Research Fellowships from the NHMRC. MS was supported by the Australian Government Research Training Program (RTP) and the Faculty of Health Scholarship at Queensland University of Technology, Australia. This study was conducted using data from UK Biobank (application number 25331), MIA (Australia), QSkin (Australia), Leeds Melanoma Cohort (UK), 23andMe Research (USA) and GWAS summary data from the melanoma meta-analysis consortium. We gratefully acknowledge Simone Cross, as well as Susan List Armitage and the Sample Processing Facility, at QIMR Berghofer Medical Research Institute for their assistance in genotyping MIA samples, and M Teresa Landi at the National Cancer Institute for the genotyping them. We would like to thank the research participants and employees of 23andMe for making this work possible.

## MELANOMA INSTITUTE AUSTRALIA

The authors thank and acknowledge the valuable contributions of Hazel Burke and Valerie Jakrot, and their colleagues, in the clinical, data management, and biospecimen banking teams at Melanoma Institute Australia. This study was supported by funding from Melanoma Institute Australia, the Australian National Health and Medical Research Council (NHMRC) through a program grant to GJM, RAS, JFT & GVL (APP1093017) and from Cancer Institute New South Wales and infrastructure grants from Macquarie University and the Australian Cancer Research Foundation. R.A.S. and G.V.L. are supported by NHMRC Fellowships (APP1141295 for R.A.S), and G.V.L. is supported by the University of Sydney Medical Foundation.

## LEEDS MELANOMA COHORT

The Leeds Melanoma Cohort was funded by the Cancer Research UK (under project grant C8216/A6129 and programme award C588/A19167), and by the National Institutes of Health (NIH) (R01 CA83115) and EU FP6 Network of Excellence award to GenoMEL. Participant recruitment was also supported by the UK National Cancer Research Network. DTB and MMI were supported in part by the Cancer Research UK awards.

