## Supplementary Information for "Higher polygenic risk for melanoma is associated with improved survival"

**Mathias Seviiri*** (ORCiD 0000-0002-8610-3283)**[1,2,3]**, **Richard A. Scolyer** (ORCiD 0000-0002-8991-0013)[4,5,6,7], **D. Timothy Bishop** (ORCiD 0000-0002-8752-8785)[8], **Mark M. Iles** (ORCiD 0000-0002-2603-6509)[9,10], **Serigne N. Lo** (ORCiD 0000-0001-5092-5544)[4,5], **Johnathan R. Stretch** [4,5,11], **Robyn P.M. Saw** [4,5,11], **Omgo E. Nieweg** [4,5,11], **Kerwin F. Shannon** [4,11,12], **Andrew J. Spillane** [4,5,13], **Scott D. Gordon** (ORCiD **0000-0001-7623-328X**)[14], **Catherine M. Olsen** (ORCiD 0000-0003-4483-1888)[15,16], **David C. Whiteman** (ORCiD 0000-0003-2563-9559)[15], **Maria T. Landi** (ORCiD 0000-0003-4507-329X)[17]**, John F. Thompson** (ORCiD 0000-0002-2816-2496)[4,5,11]**,** **Georgina V. Long (**ORCiD 0000-0001-8894-3545)[4,5,18,19], **Stuart MacGregor** (ORCiD 0000-0001-6731-8142)**[1,2]**, and **Matthew H. Law** (ORCiD 0000-0002-4303-8821)**[1,2*]**

#### Affiliations

1. Statistical Genetics Lab, QIMR Berghofer Medical Research Institute, Brisbane, QLD, Australia.
2. School of Biomedical Sciences, Faculty of Health, Queensland University of Technology, Brisbane, QLD, Australia.
3. Center for Genomics and Personalised Health, Queensland University of Technology, Brisbane, QLD, Australia.
4. Melanoma Institute Australia, The University of Sydney, Sydney, NSW, Australia.
5. Faculty of Medicine and Health, The University of Sydney, Sydney, NSW, Australia.
6. Department of Tissue Oncology and Diagnostic Pathology, Royal Prince Alfred Hospital, Sydney, NSW, Australia.
7. NSW Health Pathology, Sydney, NSW, Australia.

8. Division of Haematology and Immunology, Leeds Institute of Medical Research at St James', University of Leeds, Leeds, UK.

9. Leeds Institute of Medical Research, University of Leeds, Leeds, UK.

10. Leeds Institute of Data Analytics, University of Leeds, Leeds, UK.

11.Department of Melanoma and Surgical Oncology, Royal Prince Alfred Hospital, Camperdown, NSW Australia.

12. Sydney Head & Neck Cancer Institute, Chris O'Brien Lifehouse Cancer Center, Sydney, NSW, Australia.

13. Department of Breast and Melanoma Surgery, Royal North Shore Hospital, Sydney, NSW, Australia.

14. Genetic Epidemiology Lab, QIMR Berghofer Medical Research Institute, Brisbane, QLD, Australia.

15. Cancer Control Group, QIMR Berghofer Medical Research Institute, Brisbane, QLD, Australia.

16. Faculty of Medicine, University of Queensland, Brisbane, QLD, Australia.

17. Division of Cancer Epidemiology and Genetics, National Cancer Institute, National Institutes of Health, Bethesda, Maryland, USA.

18. Department of Medical Oncology, Mater Hospital, North Sydney, NSW, Australia.

19. Department of Medical Oncology, Royal North Shore Hospital, St Leonards, NSW, Australia.

**+61738453809,** Statistical Genetics Lab, QIMR Berghofer Medical Research Institute, 300 Herston Road, Herston QLD 4006, Australia.

#### Supplementary Methods

##### Exclusion of genetic variants associated with pigmentation and naevus pathway loci

Pigmentation and naevus count loci are major biological pathways for CM-susceptibility (Duffy et al. 2018; Landi et al. 2020). We further explored whether any PRS_susceptibility and MSS association was driven by genetic variation associated with pigmentation and/or naevi pathways. Using the previously published CM GWAS (Landi 2020), we identified both pigmentation and naevus count loci and removed SNPs from the PRS_susceptibility within +/- 0.50 megabase (mb) for each lead SNP for pigmentation (**Supplementary Table 3**) and naevi (**Supplementary Table 4)**. In addition, for loci in regions with long-range LD we excluded wider windows including *MC1R* on chromosome 16 (87-90.3 megabases), *ASIP* on chromosome 20 (30-36 megabases), and *CDKN2A* on chromosome 9 (1 megabase either side of rs871024).

Using this information we generated two additional PRS_susceptibility models; one excluding the SNPs in the pigmentation pathway ($PRS_{CMexP}$) and another excluding the loci in the naevus pathway ($PRS_{CMexN}$). First, we assessed whether new PRS models were still associated with melanoma risk using QSkin data and adjusting for covariates as before (i.e. CM risk ~ $PRS_{CMexP}$ or $PRS_{CMexN}$ + age + sex +10 PCs) (**Supplementary Figure 3**). Second, we explored if $PRS_{CMexP}$ and ($PRS_{CMexN}$ were associated with MSS in both MIA and UKB (**Results**). Third, as previously (in the **Methods**) the resulting HRs were combined by meta-analysis (**Results; Figure 2**).

##### Impact of melanoma survival prognostic factors in the MIA Cohort

Since the MIA cohort had information on other important prognostic factors including the American Joint Committee on Cancer (AJCC) 2010 stage at diagnosis (I through IV), primary tumour thickness (mm), we performed more sensitivity analyses in this dataset. First out of the 5,762 participants we excluded those with AJCC 2010 Stage O (N=90), unknown stage (N=119), and missing data (N=2) to restrict the analysis to participants with invasive melanoma (total N=5,551, melanoma deaths=771). Second, we computed Cox proportional-hazard models for MSS (MSS ~ PRS_susceptibility + 10 PCs + age + sex) (**Results**).

Next we restricted the analyses to participants with invasive melanoma and complete data on both tumour stage and primary tumour thickness (N = 5,282, melanoma deaths = 669), to compute Cox proportional-hazard models for MSS adjusting (in addition to the above) for AJCC 2010 tumour stage (stage III/IV vs stage I/II) and primary tumour thickness (thick (>1mm) vs thin (<=1mm) (MSS ~ PRS_susceptibility + 10 PCs + age + sex + tumour stage + primary tumour thickness) (**Results, Figure 3**).

To rule out survival bias due to leading time bias, we also excluded the first two years of follow up (leaving N = 4,018, and melanoma deaths = 574) and computed the Cox proportional-hazard models for MSS (MSS ~ PRS_susceptibility + 10 PCs + age + sex + tumour stage + primary tumour thickness) **(Results)**

We further examined whether there was an interaction between the $PRS_{CM}$ and strong MSS prognostic factors; by fitting interaction terms between the PRS and them and computing the Cox proportional-hazard models for MSS (MSS ~ PRS_susceptibility * tumour stage + 10 PCs + age + sex + primary tumour thickness, MSS ~ PRS_susceptibility * primary tumour thickness + 10 PCs + age + sex + tumour stage, and MSS ~ PRS_susceptibility * primary tumour thickness * tumour stage + 10 PCs + age + sex) (**Results; Table 3**).

In a similar way, we explored whether the MSS ~ $PRS_{CM}$ association differed by tumour stage (stage III/IV vs stage I/II) or primary tumour thickness (thick vs thin ) by computing the Cox proportional-hazard models for MSS in each stratum (e.g. in stage III/IV; MSS ~ PRS_susceptibility + 10 PCs + age + sex, or in thin melanoma; MSS ~ PRS_susceptibility + 10 PCs + age + sex). In order to rule whether the $PRS_{CM}$ - MSS association was not mediated through tumour or tumour thickness, we tested whether the $PRS_{CM}$ was associated with advanced tumour stage (I/II- no, and III/IV- yes) (advanced tumour stage ~ PRS_susceptibility + 10 PCs + age + sex), and primary tumour thickness (thin -no, and thick -yes) (thick tumour ~ PRS_susceptibility + 10 PCs + age + sex) (**Results; Figure 3**)

We also assessed whether the association between $PRS_{CM}$ and MSS varied across the PRS strata (quartiles) by comparing the odds of dying from melanoma for participants in the Q4, Q3, Q2 with their counterparts in Q1, adjusting for age, sex and 10 PCs using the MIA cohort (**Supplementary Figure 4**).

#### Development of the standalone skin colour polygenic risk score

We used the skin colour phenotype (data field 1717) in the UKB and coded in the order brown, dark olive, light olive, fair and very fair. Using R we applied a rank inverse normal transformation (rankit transformation) such that it could be analysed as an approximately normally distributed quantitative phenotype. We conducted a GWAS using linear mixed models using BOLT-LMM (Loh et al. 2015), adjusting for sex, age and the first ten PCs. We excluded participants with melanoma (who were included in the MSS analysis), of non-European ancestry and those who withdrew their participation in the UKB research. Following these filters, 427,893 participants were included in the analysis.

Next, we selected non-ambiguous, autosomal, bi-allelic SNPs with a minor allele frequency (MAF) > 1% and imputation quality score of 0.3 that were present in the validation (QSkin) and target (MIA and UKB) cohorts as well as the linkage disequilibrium (LD) reference panel, resulting in 6,360,404 SNPs.

Next, as for the PRS_susceptibility (**Methods**) we used LDpred (Vilhjálmsson et al. 2015) to derive PRS models at 2 mb and 5 mb of LD radii with different fractions of causal SNPs i.e. 1 (F0), 0.1 (F1), 0.01 (F2), 0.001 (F3), 0.0001 (F4) and 0.00002 (F5). We used the QSkin Cohort (1,285 melanoma cases and 15,423 controls) to validate the derived PRS models and select the best performing one. First, we used the LDpred-adjusted effect sizes (log ORs) and the imputed allelic dosages to compute PRS for each individual using PLINK 1.9 (Chang et al. 2015). Then we calculated and used Nagelkerke's $R^2$ (Nagelkerke 1991) to select the optimally performing PRS model by comparing the model fit for CM risk ~ Pigmentation PRS +age + sex +10 PCs, and a null model (CM risk ~ age + sex +10 PCs). Model performances are presented in **Supplementary Figure 2**. The best performing PRS model was subsequently used to explore the association between the skin colour PRS and melanoma risk in the QSkin cohort, and melanoma specific survival in UKB and MIA cohorts. The F1 2 mb LD radius model was the best performing model with Nagelkerke's $R^2$ of 3.1% (**Supplementary Figure** 2). It was thus used in all our subsequent analyses.

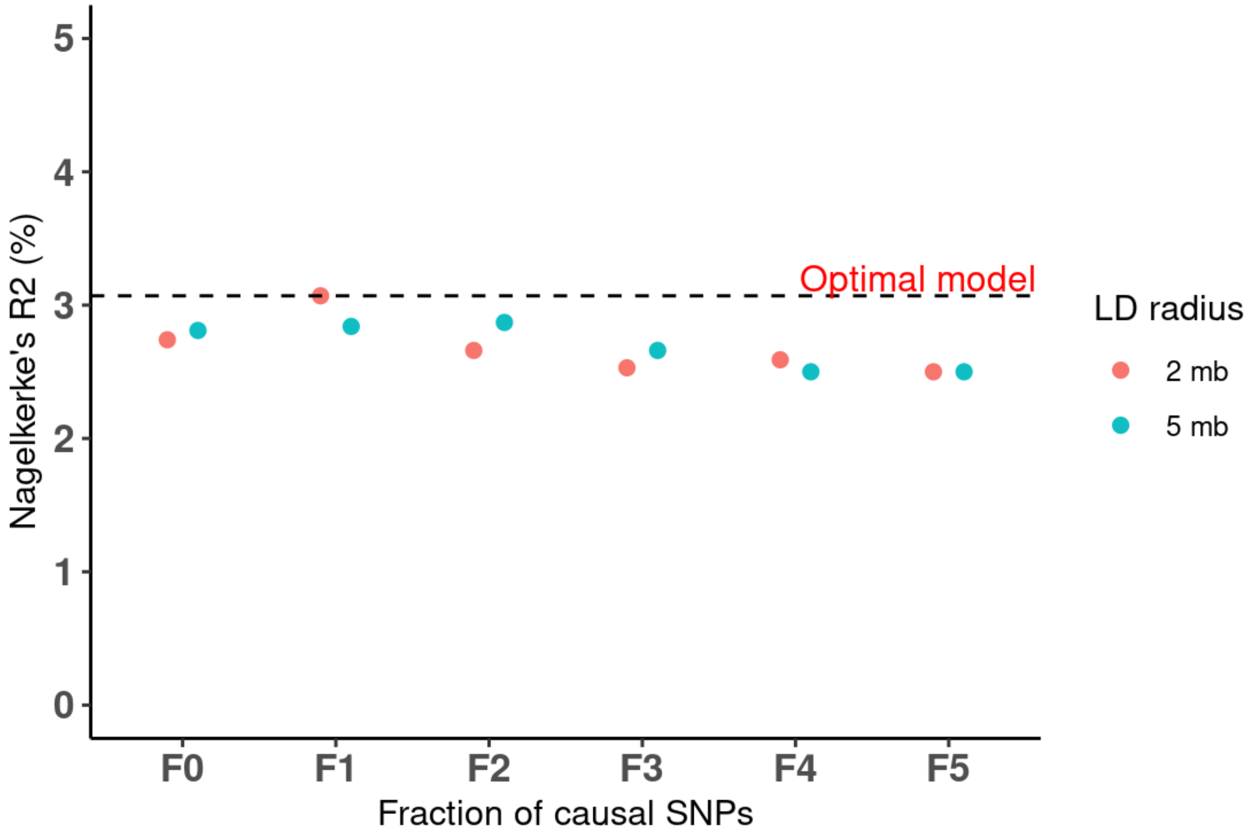


**Supplementary Figure 2: Skin colour polygenic risk score model performance in the validation cohort (QSkin).** *The x-axis represents the different melanoma* polygenic risk score (PRS) *models of varying fractions of causal SNPs (i.e. 1 (F0), 0.1 (F1), 0.01 (F2),0.001(F3),0.0001 (F4) and 0.00002 (F5)) at the different radii of the linkage disequilibrium (LD) (i.e. 2 megabase (mb) and 5 mb). The y-axis represents Nagelkerke's* $R^2$ *(%) for each of the 12 PRS models. The horizontal dashed black line highlights the optimal model (F1-2mb) (i.e. with the highest Nagelkerke's* $R^2$*).*

#### Generation of the standalone naevus count pathway polygenic risk score

We used naevus count PRS (**Supplementary Table 5**) of genome wide significant ($5x10^{-8}$) SNPs from a previously published GWAS of naevus count (Duffy et al. 2018). In brief, the GWAS included 52,506 participants of European ancestry without melanoma, non-overlapping with the QSkin, MIA or UKB cohorts, from 11 cohorts from Europe, Australia, and USA. Details of the included cohorts and other quality control metrics have been published elsewhere (Duffy et al. 2018). Individual scores were generated in QSkin, MIA and UKB by using the SNP effect sizes (betas) as the weights and the imputed allelic dosages using PLINK 1.9 and analysed as done for the pigmentation PRS above.

#### Development and assessment of the standalone telomere length polygenic risk score.

Using data from the UKB we conducted a GWAS on telomere length (data field 22192; Z-adjusted T/S log) using linear mixed models using BOLT-LMM v2.3 (Loh et al. 2015), adjusting for sex, age and the first ten PCs. For the telomere length phenotype, adjusted leukocyte telomere length (Field 22191) was both loge-transformed to obtain a normal distribution and then Z-standardised (https://biobank.ndph.ox.ac.uk/ukb/field.cgi?id=22192). After excluding all participants who were included in the MSS analysis, of non-European ancestry and those who withdrew their participation in the UKB research, 433,431 individuals were available for the analysis. After the conducting the GWAS, we retained non-ambiguous, autosomal, bi-allelic SNPs with a minor allele frequency (MAF) > 1% and imputation quality score of 0.3 which overlapped in the validation (QSkin) and target (MIA and UKB) cohorts.

After performing LD clumping ($r^2 = 0.5\%$, and 5000 kb, and $P < 1$) to select independent SNPs, we generated 8 PRS models based on P-value thresholds less than; $5 \times 10^{-8}$, $10^{-7}$, $10^{-6}$, $10^{-5}$, $10^{-4}$, $10^{-3}$, $10^{-2}$, and $10^{-1}$ and validated them in the QSkin Cohort (as described for skin colour PRS) to select the optimal telomere length PRS ($PRS_{TL}$). PLINK/1.90b6.8 (Chang et al. 2015) was used for LD clumping. The PRS at $P < 5 \times 10^{-8}$ was selected as the best performing one (Nagelkerke's $R^2$ of 2.6%). First, we tested if $PRS_{TL}$ was associated with CM risk in the QSkin cohort (**Supplementary Figure 3)**, then with MSS in the MIA cohort (**Results**).

#### Testing the association between skin colour, naevus count and telomere length genetics and melanoma specific survival

In addition, we assessed if independent polygenic risk scores for pigmentation ($PRS_P$), naevus count ($PRS_N$) and telomere length ($PRS_{TL}$) were associated with MSS. First, we generated $PRS_P$, $PRS_N$, and $PRS_{TL}$ using data independent of the QSkin (validation cohort) and selected the best models as described previously. After validating that these PRS were associated with CM risk in QSkin (**Supplementary Figure 3**), we tested if they were associated with MSS in UKB and MIA, as above; MIA and UKB estimates were combined by fixed effects meta-analysis (**Supplementary Figure 5 (**$PRS_P$ & $PRS_N$**)** and **Results (**$PRS_{TL)}$ )).

### Supplementary Results

#### Supplementary Figure 1:

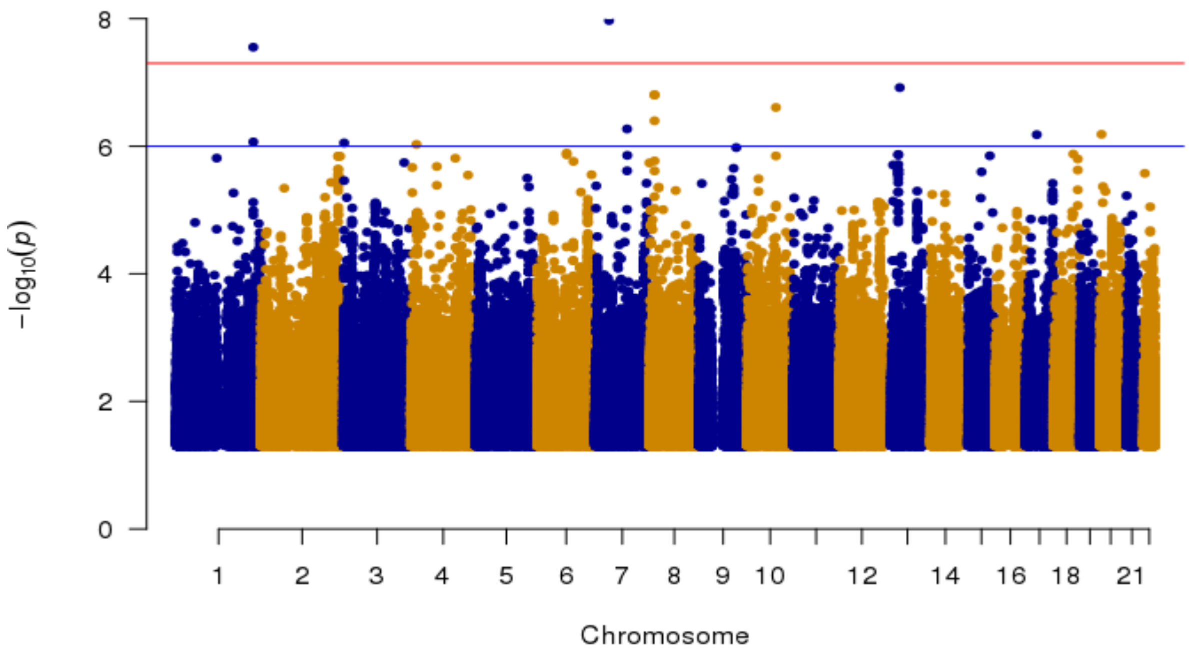


**Supplementary Figure 1:** Manhattan plot for the MSS GWAS meta-analysis between MIA and UKB cohorts.

**Association of the cutaneous melanoma polygenic risk score with melanoma risk.**

After adjusting for age at diagnosis, sex and the first ten PCs, a one SD increase in the $PRS_{CM}$ was positively associated with CM risk (OR = 1.80, 95% CI = 1.69—1.92, P = 8.7 × $10^{-71}$) in the validation cohort (**Supplementary Figure 3**). The association was largely contributed, but not entirely driven by the pigmentation and naevus pathways. The $PRS_{CM}$ models without the naevus and pigmentation genomic regions were still associated with increased risk of CM ($PRS_{\mathbf{CMexN}}$ ; OR = 1.53, 95% CI = 1.43—1.64, P = 3.99 × $10^{-37}$ and $PRS_{\mathbf{CMexP}}$; OR = 1.35, 1.27—1.43, P = 2.46 × $10^{-24}$). The naevus count, skin colour (pigmentation) and telomere length PRSs were also associated with increased risk melanoma ($PRS_N$; OR = 1.16, 95% CI = 1.09—1.22, P = 6.32 × $10^{-7}$, $PRS_{\mathbf{P}}$; OR = 1.22, 95% CI = 1.15—1.30, P = 7.58 × $10^{-11}$, and $PRS_{TL}$; OR = 1.38, 95% CI= 1.05—1.82, P = 0.0230).

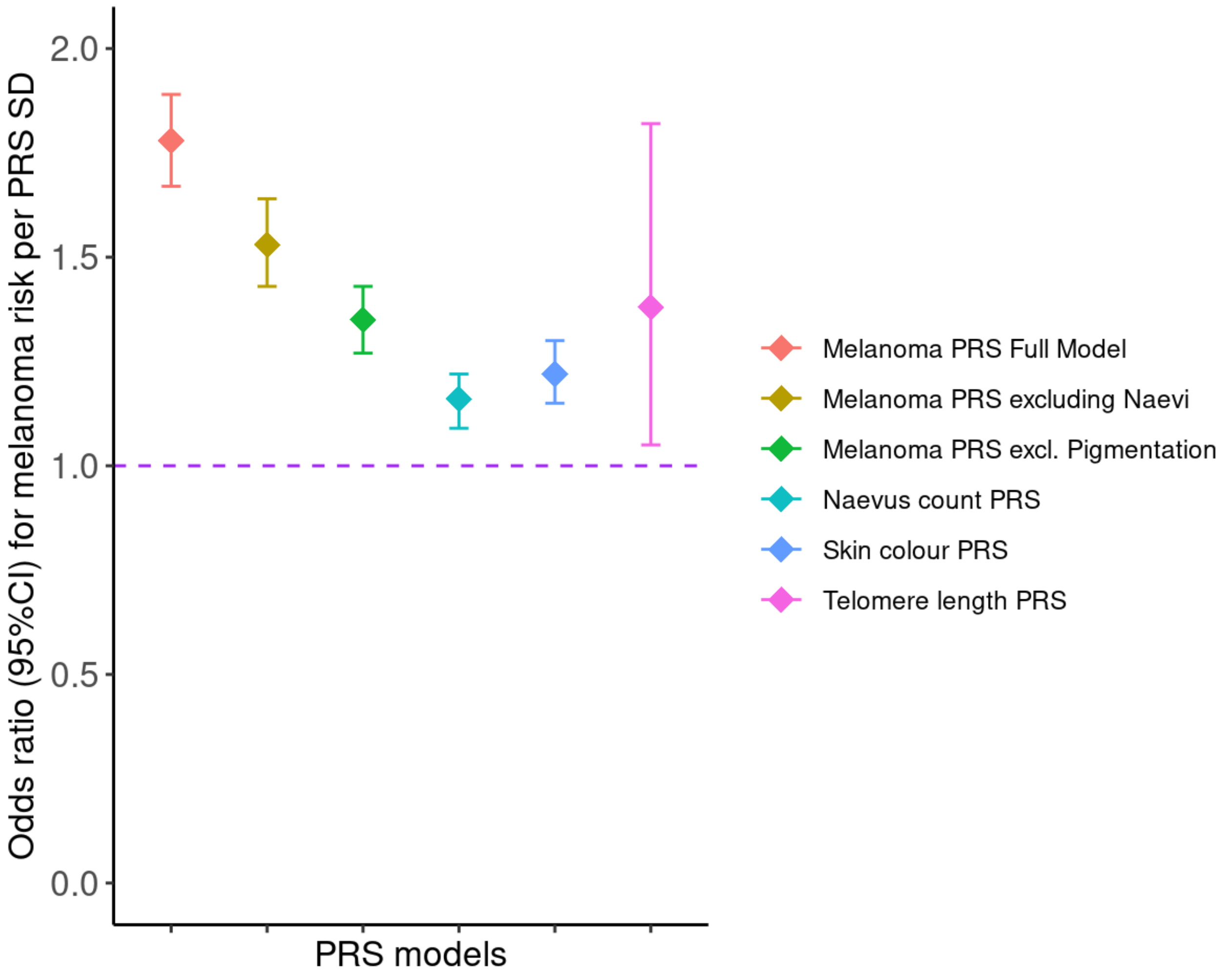

***Supplementary Figure 3: The association between polygenic risk scores and the risk of melanoma in QSkin.***

*The x-axis represents the different polygenic risk score (PRS) models represented in red (Full melanoma model), dark green (melanoma model after excluding the naevus count loci), green (melanoma model after excluding the pigmentation loci), cyan (an independent naevus count PRS model), blue (an independent skin colour PRS) and pink (an independent telomere PRS). Skin colour phenotype was rank normalised in the order brown, dark olive, light olive, fair and very fair. The y-axis represents the odds ratio for melanoma risk per standard deviation (SD) increase in the respective PRSs. Error bars are the 95% confidence interval for each PRS model. All models were adjusted for age, sex and the first ten principal components.*

#### Melanoma prognostic factors, PRS and survival

The PRS_susceptibility was also not a predictor of advanced tumour stage (I/II- no, and III/IV- yes) (OR =1.02 95%CI= 0.94- 1.10, P= 0.64357) and primary tumour thickness (thin -no, and thick -yes,) (OR = 1.003, 95%CI= 0.95-1.06, P= 0.89580). Yet advanced tumour stage and primary tumour thickness (as expected) were strong predictive factors for MSS (advanced stage; HR =3.12, 95%CI= 2.63 - 3.70, P = 5.97 x$10^{-40}$; adjusted for age, sex, tumour thickness and 10 PCs, and tumour thickness (thick vs thin); HR = 2.00, 95%CI=1.63- 2.47, P= 6.93 x $10^{-11}$).

##### Variation of MSS across PRS quartile in the MIA cohort

Compared to the bottom quartile (Q1), participants in the top quartile (Q4) with the highest genetic risk for CM were 38% less likely to die due to melanoma (HR = 0.62, 95% CI = 0.51-0.76, *P* = 2.34 × $10^{-6}$) (**Supplementary Figure 4**).

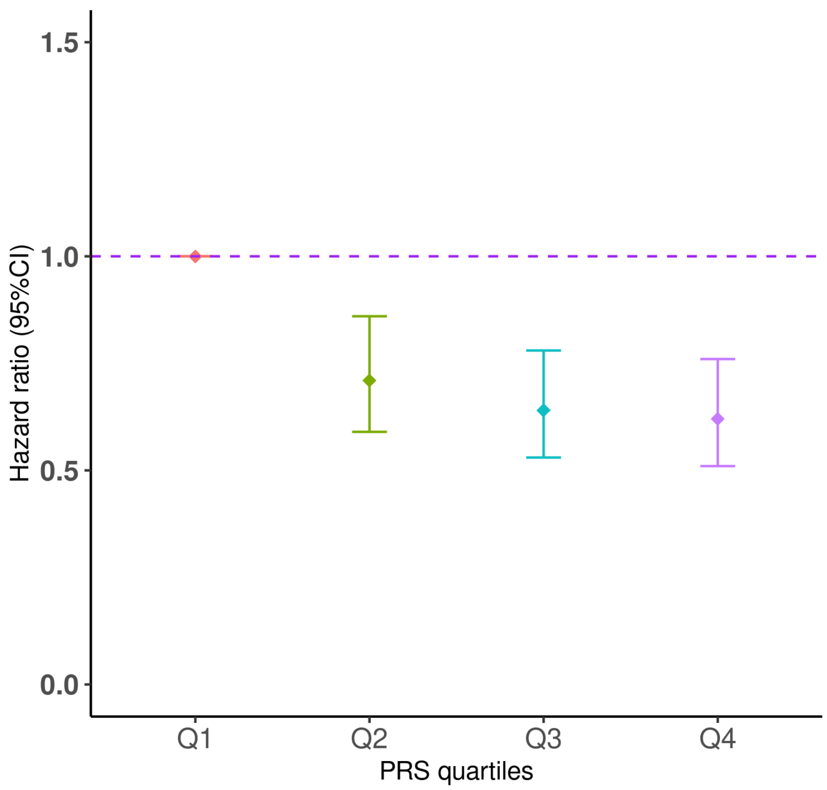


***Supplementary Figure 4: The association by quartile of polygenic risk for melanoma susceptibility and melanoma specific survival in the MIA cohort***

*This analysis of the MIA cohort includes* 5,762 patients, of which 800 died from melanoma. *The x-axis represents the different quartiles for the CM polygenic risk score (PRS) from Q1 to Q4. The y-axis represents the hazard ratio (HR) and the 95% confidence interval (CI) computed using Cox proportional-hazards models for each quartile adjusting for age at diagnosis, sex, 10 PCs and genotype batch effects.*

**Supplementary Figure 5: Association of standalone skin colour and naevus PRSs and melanoma specific survival in MIA and UKB.**

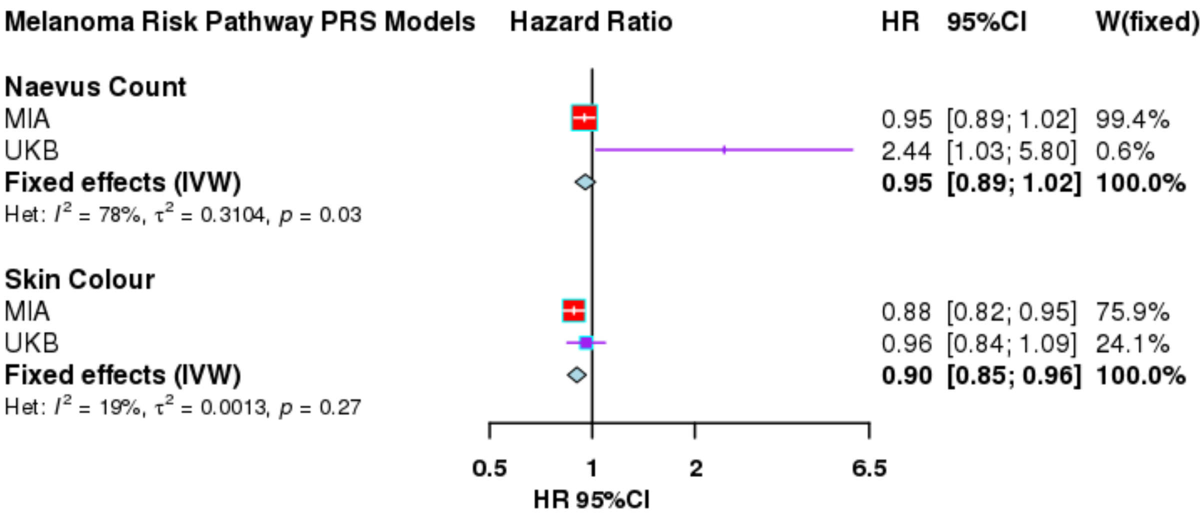

### SUPPLEMENTARY REFERENCES

Chang, Christopher C., Carson C. Chow, Laurent Cam Tellier, Shashaank Vattikuti, Shaun M. Purcell, and James J. Lee. 2015. "Second-Generation PLINK: Rising to the Challenge of Larger and Richer Datasets." *GigaScience* 4 (February): 7.

Duffy, David L., Gu Zhu, Xin Li, Marianna Sanna, Mark M. Iles, Leonie C. Jacobs, David M. Evans, et al. 2018. "Novel Pleiotropic Risk Loci for Melanoma and Nevus Density Implicate Multiple Biological Pathways." *Nature Communications* 9 (1): 4774.

Landi, Maria Teresa, D. Timothy Bishop, Stuart MacGregor, Mitchell J. Machiela, Alexander J. Stratigos, Paola Ghiorzo, Myriam Brossard, et al. 2020. "Genome-Wide Association Meta-Analyses Combining Multiple Risk Phenotypes Provide Insights into the Genetic Architecture of Cutaneous Melanoma Susceptibility." *Nature Genetics* 52 (5): 494–504.

Loh, Po-Ru, George Tucker, Brendan K. Bulik-Sullivan, Bjarni J. Vilhjálmsson, Hilary K. Finucane, Rany M. Salem, Daniel I. Chasman, et al. 2015. "Efficient Bayesian Mixed-Model Analysis Increases Association Power in Large Cohorts." *Nature Genetics* 47 (3): 284–90.

Nagelkerke, N. J. D. 1991. "A Note on a General Definition of the Coefficient of Determination." *Biometrika* 78 (3): 691–92.

Vilhjálmsson, Bjarni J., Jian Yang, Hilary K. Finucane, Alexander Gusev, Sara Lindström, Stephan Ripke, Giulio Genovese, et al. 2015. "Modeling Linkage Disequilibrium Increases Accuracy of Polygenic Risk Scores." *American Journal of Human Genetics* 97 (4): 576–92.
